# National consensus statement on opioid agonist treatment in custodial settings

**DOI:** 10.1101/2024.08.15.24312029

**Authors:** Jocelyn Chan, Jon Cook, Michael Curtis, Adrian Dunlop, Ele Morrison, Suzanne Nielsen, Rebecca Winter, Thileepan Naren, the National Prisons Addiction Medicine Network

## Abstract

**Introduction:** Opioid use and dependence are prevalent among incarcerated people, contributing to elevated rates of overdose and other harms in this population. Opioid agonist treatment (OAT) has been demonstrated as an effective intervention to mitigate these risks. However, challenges to health care implementation in the custodial sector result in suboptimal and variable access to OAT in prisons nationally.

**Main recommendations:** Among a national multi-disciplinary expert panel, we conducted a modified Delphi study which yielded 19 recommendations to government, relevant health authorities and custodial health services. These recommendations cover five core domains: induction or continuation of OAT, OAT options and administration, transition of care to the community, special populations, organisational support. Key recommendations include prompt recognition and treatment of opioid withdrawal, active linkage to community-based OAT providers upon release, and ensuring appropriate organisational support through local protocols, adequate funding, and monitoring of key program indicators.

**Changes in management as a result of this statement:** This consensus statement addresses a significant gap in national policy on OAT in Australian prisons. The recommendations set forth best practice standards grounded in evidence and expert consensus. We expect that implementing these recommendations will enhance the quality, consistency, and continuity of OAT both within prison and upon release. Optimizing OAT provision is crucial for improving health outcomes and addressing overdose, which is the leading cause of death among people released from prison.

## Background

In Australia, more than 40 000 people are incarcerated on any given day. (1) Among incarcerated people, the prevalence of substance use and substance dependence is high – in large part due to the criminalisation of drug use. (2) In Australia, surveys indicate almost two-thirds of prison entrants had used illicit drugs in the previous year, over 50% of people who are incarcerated have a substance dependence and over 30% have an opioid dependence. (2–4)

Opioid agonist treatment (OAT) is an effective and evidence-based treatment for opioid dependence. (5) Access to and retention in OAT reduces drug-related harms, including blood-borne virus acquisition and overdose, in prison and on release. (6) However, access to and uptake of OAT in prisons is suboptimal and inconsistent. (7) Access to OAT in custodial settings varies across Australia; custodial healthcare is overseen by jurisdictional authorities and delivered through a diverse array of both public and private healthcare providers. Barriers to providing OAT in custodial settings include jurisdictional or institutional policies that restrict provision of OAT, limited capacity or resources for provision of OAT, and negative societal attitudes to OAT.(8) Access to health care in short-term custody settings, including police cells or watch-houses, can be particularly challenging. (9)

The National Prisons Addiction Medicine Network was convened to address a gap in the national policy landscape relating to the provision of evidence-based best-practice medical care for incarcerated people with substance dependence, including OAT. The network comprises clinical, consumer and public health stakeholders from a range of jurisdictions with relevant experience in providing addiction and broader health services to incarcerated people.

This consensus statement aims to improve quality, consistency, and continuity of OAT for people who are incarcerated in Australia by promoting a nationally coordinated and evidence-based approach to OAT provision and identifying targets against which to monitor progress. In this statement, we adhere to the principles outlined in Rule 24 of the United Nations Standard Minimum Rules for the Treatment of Prisoners (the Mandela Rules) stipulating that prisoners are entitled to medical care that is equivalent to that which they could access in the community. This statement has been developed for application to all custodial settings, including adult and juvenile prisons, jails and police cells (or watch-houses). The objectives are:

- to present a critical analysis of the evidence supporting the provision of OAT in custodial settings, and
- to develop consensus recommendations for the provision of OAT in custodial settings.

## Methods

### Expert panel and scope

An expert panel was convened to represent a broad spectrum of expertise in the fields of addiction medicine and custodial health. (Table 1) This panel is comprised of 18 clinical, consumer and public health stakeholders representing all Australian jurisdictions. (Appendix 1) An initial meeting of the expert panel was held to review and agree upon the scope of the consensus statement (6 Nov 2023).

**Table 1:**
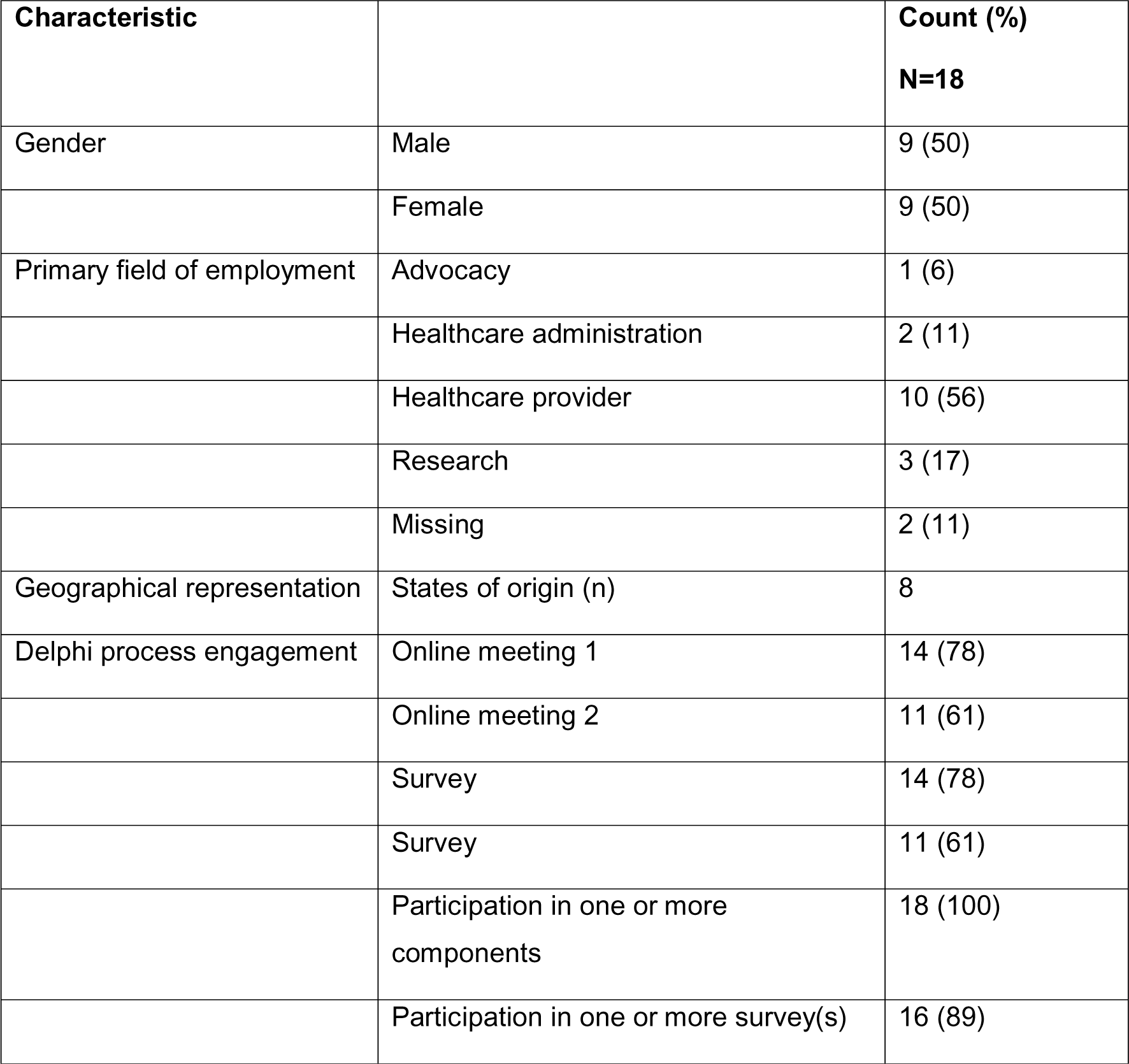
Expert panel demographic composition and level of engagement.

### Literature review

Published literature from January 2002 to November 2023 was searched. Relevant papers were identified by searching PubMed using the key words “methadone” or “buprenorphine” or “opioid substitution treatment” and “prisons” or “prisoners” or “correctional facilities”. Additional relevant papers were nominated by expert panel members. Original research, including systematic reviews, randomised controlled trials observational studies and qualitative studies, relating to the provision of OAT in prisons, including transition from incarceration to the community, were reviewed.

### Modified Delphi methods

The research team drew on the literature identified in the reviews to develop a draft set of recommendations. The expert panel provided input on the draft recommendations via an online meeting (8 Feb 2024) and email feedback (response period from 8 Feb-12 Mar 2024). The recommendations were further refined through two rounds of surveys (R1 in May 2024, R2 in July 2024), developed and distributed using the Qualtrics XM platform. Both surveys contained 19 recommendations and response options to indicate agreement (agree *vs* disagree). In the first survey, comments for each recommendation were collected through a free text entry option. Consensus was defined a priori as a greater than 80% agreement from respondents. Recommendations that reached consensus were included in the final document. The survey responses were collated to grade each recommendation based on the level of agreement in which ‘U’ denotes unanimous (100%) agreement, ‘A’ 90-99% agreement and ‘B’ 80-89% agreement. Methods were based on recently published consensus statements. (10–12)

### Consensus recommendations

Among a national multi-disciplinary expert panel, the Delphi process yielded 19 recommendations for the provision of OAT in custodial settings in Australia. Fourteen recommendations achieved unanimous agreement (U) (Table 2).

**Table 2:** Consensus recommendations.

| 1 | Induction or continuation of OAT | Grade* |
| --- | --- | --- |
|  | We recommend that custodial health services: |  |
| 1.1 | Continue treatment for people entering custodial settings on OAT without interruption. | U |
| 1.2 | Confidentially screen people entering custodial settings for opioid dependence and risk of opioid withdrawal. | U |
| 1.3 | Assess and treat people at risk of opioid withdrawal within 24 hours. They should be monitored by appropriately qualified health care providers for at least 72 hours following detention. | U |
| 1.4 | Offer OAT to all who meet criteria for opioid dependence according to International Classification of Diseases, 11<sup>th</sup> Revision (ICD-11) or Diagnostic and Statistical Manual of Mental Disorders, Fifth Edition (moderate-severe opioid use disorder, DSM-5). The principles of informed consent should be observed. There should be no arbitrary limits to OAT access based on resource constraints. | U |
| 1.5 | Offer a health assessment to people seeking OAT at any time during their incarceration, within two weeks. Earlier assessment is required for people at risk of opioid withdrawal (see recommendation 1.3). Priority should be given to pregnant women and people with significant physical or mental health comorbidities. | A |
| **2** | **OAT options and administration** |  |
|  | We recommend custodial health services: |  |
| 2.1 | Use a person-centred approach that allows choice of medication. The choice of medication and formulations offered is a clinical decision that requires thorough consideration of the risks and benefits for each individual. | B |
| 2.2 | Consider maximising access to the long-acting buprenorphine depot, given it may facilitate greater treatment access with the same resources. | A |
| 2.3 | Avoid withholding or discontinuing OAT as a disciplinary measure. Forced tapering and withdrawal of OAT during incarceration increases risk of overdose and death on release. | A |
| **3** | **Transition of care to the community** |  |
|  | We recommend that custodial health services: |  |
| 3.1 | Actively link people on OAT with community-based OAT providers prior to release to facilitate continuity of care. | U |
| 3.2 | Provide individuals with a bridging prescription and accessible dosing location on release from prison. The script should be of sufficient duration to ensure continuity of treatment while identifying a community prescriber – ideally at least four weeks supply. Ensure that take-away doses are available | U |
|  | for days when pharmacies are closed and supervised dosing is not available. |  |
| 3.3 | Implement programs to provide psychosocial support on release, including peer or patient navigators, to improve OAT retention. | A |
| 3.4 | Provide training and access to take-home naloxone during incarceration and on release to reduce the risk of fatal overdose. | U |
| **4** | **Special populations** |  |
|  | We recommend that custodial health services: |  |
| 4.1 | Collaborate with Aboriginal and Torres Strait Islander community representatives, including Elders, to ensure culturally appropriate care for Aboriginal and Torres Strait Islander people. Consider establishment of in-reach services in collaboration with local Aboriginal Community Controlled Health Organisations. | U |
| 4.2 | Continue or commence OAT for pregnant women with opioid dependence. | U |
| **5** | **Organisational support** |  |
|  | We recommend that custodial health services: |  |
| 5.1 | Maintain up-to-date protocols or guidelines for OAT service delivery. | U |
| 5.2 | Implement opioid harm reduction education programs, covering OAT, overdose prevention and stigma, for people in prison, healthcare providers and correctional staff. The program should be culturally appropriate and accessible to people with varying levels of health literacy. | U |
|  | We recommend that government and relevant health authorities: |  |
| 5.3 | Ensure adequate and sustained funding to support OAT service delivery. | U |
| 5.4 | Implement a jurisdiction-wide electronic medical record in custodial settings to promote continuity of care across settings. | U |
| 5.5 | Monitor key OAT program indicators, including screening, uptake, wait-times, retention, and adverse events to inform ongoing quality improvement. | U |

\* Grading of consensus responses: 'U' denotes unanimous (100%) agreement, 'A' 90-99% agreement and 'B' 80-89% agreement.

### Supporting literature

#### Induction or continuation of OAT

The provision of OAT is associated with a range of positive outcomes for individuals with opioid dependence both during incarceration and in the high-risk period immediately post-release. There is consistent evidence for significant reductions in illicit opioid use, injecting drug use and syringe sharing during imprisonment from systematic reviews of trials and observational studies. (13, 14) Systematic reviews of trials indicate provision of OAT in prison, particularly methadone, improves post-release outcomes including decreasing mortality, increasing engagement with community-based treatment, reducing opioid use and injecting drug use. (13, 15, 16)

Given the high prevalence of opioid dependence and opioid related harms among incarcerated people and the availability of an effective treatment, we recommend screening for opioid dependence among all people entering custodial settings. Evidence suggests that screening for substance dependence is feasible and reliable in the prison setting. (17–19) Providing timely access to adequate doses of OAT is important to prevent and manage opioid withdrawal. Importantly, persistent vomiting and diarrhoea can lead to severe fluid loss and electrolyte abnormalities that result in haemodynamic instability and in rare instances, death. (20) Delays in accessing OAT in custodial settings may also affect subsequent uptake of OAT. (7)

In addition to screening on entry, individuals should be provided with the opportunity to engage with treatment throughout their period of incarceration. Evidence indicates that, while prohibited, illicit drug use is common in prison settings. (21) Furthermore, for individuals with a history of injecting drug use, rapid resumption of injecting drug use following prison release is common. (22) This resumption in drug use is associated with a high risk of mortality—attributable primarily to opioid overdose – particularly in the first two weeks following release. (23, 24) Evidence supports initiation of OAT in custodial settings rather than initiation at or after release. (13, 25) In addition to evidence for positive outcomes during imprisonment, initiation in prison improves treatment retention and reduces illicit opioid use post-release compared to initiation at release or counselling only. (26)

### OAT options and administration

Evidence on the relative benefits of different OAT options in custodial settings are limited. Three small RCTs directly compared sublingual buprenorphine to methadone in prison. They reported improved treatment retention and fewer side effects with sublingual buprenorphine, but comparable clinical effectiveness in reducing illicit opioid use. (27) Preliminary evidence for the relatively newer long-acting buprenorphine depot indicates that it is safe, acceptable and lower-cost compared to other treatment options in prison. (28)

We support using a patient-centred approach that allows choice of OAT in custody. Person-centred care and shared decision-making are recommended in standards and guidelines, both locally and internationally. (29, 30) While there is limited research on how patient preference affects outcomes of OAT treatment for incarcerated people, systematic reviews from the broader mental health literature indicate incorporating patient preference improves treatment engagement and patient satisfaction, with mixed results (positive or no effect) for treatment outcomes (31). Qualitative research indicates that many people who are incarcerated prefer OAT options based on their effectiveness at treating cravings, route of delivery, side effects and structures of medication delivery in the community. (32) People have also described switching OAT options to mitigate risk of violence from peers to divert their prescribed OAT. (7)

Restrictions to the availability of OAT options in prisons are often decided based on risk of diversion or non-medical use. Diversion refers to the selling, trading, sharing or giving away of medications to recipients other than the prescribed person. (33) Evidence indicates that both methadone and buprenorphine are diverted in prison, however data on rates of diversion are scarce and available evidence indicates that patterns of non-prescribed pharmaceutical opioid use in prison are heterogenous dependent on location and context. (34) The few studies that have investigated the relative risk of diversion of different formulations in custodial settings indicate sublingual buprenorphine is more commonly diverted than methadone. (35) The long-acting buprenorphine depot, administered subcutaneously weekly or monthly, has been suggested as a potential solution to limit potential for diversion. (28) No diversion was self-reported from a recent Australian trial of the buprenorphine depot. (28)

In addition to the reduced risk of diversion and improved safety profile, the long-acting buprenorphine depot is preferred in the prison setting due to its longer dosing interval. Monthly dosing enables a period of coverage post-release, which is a time of high overdose and mortality risk. This may be particularly important for individuals on remand, who do not benefit from post-release planning. Furthermore, monthly rather than daily dosing reduces the resources required for OAT delivery, increasing capacity to enrol more individuals in the program – potentially reducing waitlists and improving access. (36–38)

Importantly, we recommend against the discontinuation of OAT or dose reduction as a disciplinary measure. (39) Forced withdrawal increases the risk of substance use during incarceration and decreases treatment engagement on release, thereby increasing the risk of death. (40, 41) People who use drugs may also be less likely to initiate OAT in the community for fear of losing access during periods of incarceration and undergoing forced withdrawal. (42)

### Transition of care to the community

The period post-release from custodial settings is characterised by a markedly increased risk of mortality, especially from opioid overdose. (23, 43, 44) Retention in OAT following release from custodial settings mitigates the risk of opioid overdose by maintaining opioid tolerance and reducing demand for illicit opioids. Retention in OAT also improves rates of primary health contact and reduces rates of ambulance or emergency department use. (45, 46) However, discontinuation from OAT following release from prison is common. (47, 48)

Peer or patient navigators have the potential to support OAT retention by helping navigate complex healthcare and social services systems. While there is strong evidence for patient navigation across a range of health domains, (49, 50) there is limited and mixed evidence on the effect of patient navigation on OAT retention post-release from prison. (51, 52) Qualitative studies support the role of peer and patient navigators post-release. (53–55)

A systematic review of qualitative evaluations of prison release programs identified factors associated in program success including access to structural supports, particularly housing and employment, and continuity of care, through fostering the formation and maintenance of a therapeutic relationship throughout the pre-release and post-release period. (56)

Given the high risk of overdose mortality after release from prison, we also recommend the provision of take-home naloxone prior to or on release, alongside training in how to use naloxone, and information about where to receive additional training and resources in the community. An evaluation of a national prison-based take home naloxone program using observational data in Scotland reported a continuous decrease in overdose-related mortality within the 4 weeks post release. (57, 58) Studies also indicate widespread support for naloxone training in custody and distribution at release among both people recently released from prison and key prison stakeholders. (59, 60)

### Special populations

Literature on OAT for incarcerated Aboriginal and Torres Strait Islander people and pregnant women was reviewed. We were unable to identify evidence related to culturally and linguistically diverse populations or youth populations.

Aboriginal and Torres strait Islander people are disproportionately incarcerated in Australia. Despite making up only 4% of the Australian population, Aboriginal and Torres Strait Islander people make up 33% of all people in prison. (61) Furthermore, Aboriginal and Torres Strait Islander people are at increased risk of OAT discontinuation on release from prison. (47)

Aboriginal and Torres Strait Islander communities have a strong history of providing holistic, culturally appropriate health care and leading responses to reduce drug-related harms. (62) In line with the United Nations Declaration on the Rights of Indigenous Peoples principle of self-determination, we recommend collaborating with Aboriginal and Torres Strait Islander communities to ensure optimal OAT treatment outcomes. (63) The Winnunga Holistic Health Care Prison Model (Australian Capital Territory) and the South Australian Prison Health Service Model of Care for Aboriginal Health and Wellbeing provide examples of models of prison health care developed in consultation with community. (64, 65) Facilitating ‘in-reach’ services from Aboriginal Community Controlled Health Organisations (ACCHOs) may improve health care delivery to Aboriginal and Torres Strait Islander people in prison and facilitate continuity of care on-release. (66) However, OAT programs are rare among ACCHOs. (67)

Women make up a small but increasing proportion of the Australian prison population. The majority of incarcerated women are of child-bearing age and in 2021, 7% of women in prison were pregnant. (68) Prevalence of substance use is high among incarcerated women, including pregnant incarcerated women. (69)

Opioid agonist treatment is strongly recommended for pregnant women with opioid dependence. In addition to reductions in opioid use and risk of opioid overdose, systematic reviews of observational studies indicate treatment with methadone or buprenorphine (including buprenorphine-naloxone) during pregnancy is associated with improved adherence to antenatal care and lower incidence of preterm birth and stillbirth. (70–72) Neither methadone nor buprenorphine appear to be teratogenic, however pregnant women need to be counselled about the risks of neonatal opioid withdrawal syndrome. (73) Within prison contexts, Peeler et al. recommend screening women for both pregnancy and opioid dependence upon intake to ensure that women receive timely treatment and avoid experiencing withdrawal. (74) This is supported by data from Australia indicating that most (88%) incarcerated women described their current pregnancy as unplanned and half were unaware they were pregnant before incarceration. (75)

### Organisational support

While there is strong evidence for the effectiveness of OAT against a range of health outcomes, in-prison coverage remains suboptimal. The National Opioid Pharmacotherapy Statistics Annual Data collection report substantial differences in the pattern of OAT prescribing by state, indicating inconsistencies in service availability and provision. (76) While quantitative estimates of unmet demand are not available, qualitative data indicates there are delays and challenges in accessing OAT due to OAT prison policy and service delivery limitations. (7)

There have been two recent reviews of qualitative studies examining the barriers to implementation of OAT within prison settings. (8, 77) Common barriers identified by both reviews included: stigma associated with OAT, a lack of knowledge about benefits of OAT among prison stakeholders, preference for abstinence-oriented treatment, lack of resources including qualified staff, a lack of appropriate policies and protocols and poor continuity of care on transfer or post-release. (8, 77) Grella et al. highlighted the inter-related nature of these barriers. For example, the societal stigma associated with OAT leads to unfavourable institutional policies, limited resourcing and resultant poor experiences (including forced withdrawals). These poor experience then reinforcing negative attitudes towards OAT among incarcerated individuals. (77)

There have been comparatively fewer studies examining facilitators to implementation of OAT in prison settings. (77) Grella et al. reviewed three intervention studies, indicating that training can improve knowledge among custodial staff as measured by surveys pre-and post-intervention, however an intervention which linked healthcare providers demonstrated greater improvements in staff attitudes and referral intentions compared to information provision alone. (77)

## Conclusion

This consensus statement, developed by a national multidisciplinary expert panel with robust representation from custodial health practitioners and supported by academic and consumer input, offers evidence-based and actionable recommendations to enhance OAT provision in custodial settings. In alignment with the Mandela Rules, the recommendations aim to ensure that incarcerated individuals receive treatment equivalent to that available in the community. Through improvement in OAT provision, both in prison and post-release, we anticipate significant gains in health outcomes and a reduction in post-release morbidity and mortality among this highly marginalised population.

## Data Availability

N/A

## Acknowledgements

The Consensus statement was prepared on behalf of the National Prisons Addiction Medicine Network (NPAMN). We acknowledge the time and contributions from everyone on the expert panel: Adrian Dunlop, Andrew Wiley, Bianca Davidde, Christine Watson, David Onu, Ele Morrison, Jeremy Hayllar, Jocelyn Chan, Katerina Lagios, Kevin Fontana, Mark Stoove, Peter Thompson, Rebecca Li, Rebecca Winter, Shalini Arunogiri, Suzanne Nielsen, Thileepan Naren, and Tom Turnbull.

The author(s) received no financial support for the research, authorship, and/or publication of this article.

## References

1. Australian Bureau of Statistics. Prisoners in Australia. Canberra: ABS; 2022 28 December 2023.

2. Butler T, Indig D, Allnutt S, Mamoon H. Co-occurring mental illness and substance use disorder among Australian prisoners. Drug and Alcohol Review. 2011;30(2):188–94.

3. Ogloff JR, Lemphers A, Dwyer C. Dual diagnosis in an Australian forensic psychiatric hospital: prevalence and implications for services. Behav Sci Law. 2004;22(4):543–62.

4. Australian Institute of Health and Welfare (AIHW). The health of Australia’s prisoners 2018. Canberra; 2019. Contract No.: Cat. no. PHE 246.

5. Strang J, Volkow ND, Degenhardt L, Hickman M, Johnson K, Koob GF, et al. Opioid use disorder. Nature Reviews Disease Primers. 2020;6(1):3.

6. Malta M, Varatharajan T, Russell C, Pang M, Bonato S, Fischer B. Opioid-related treatment, interventions, and outcomes among incarcerated persons: A systematic review. PLOS Medicine. 2019;16(12):e1003002.

7. Marshall AD, Schroeder SE, Lafferty L, Drysdale K, Baldry E, Stoové M, et al. Perceived access to opioid agonist treatment in prison among people with a history of injection drug use: A qualitative study. J Subst Use Addict Treat. 2023;150:209066.

8. Komalasari R, Wilson S, Haw S. A systematic review of qualitative evidence on barriers to and facilitators of the implementation of opioid agonist treatment (OAT) programmes in prisons. Int J Drug Policy. 2021;87:102978.

9. Crilly JL, Brandenburg C, Kinner SA, Heffernan E, Byrnes J, Lincoln C, et al. Health care in police watch-houses: a challenge and an opportunity. Med J Aust. 2022;217(6):287–9.

10. Lazarus JV, Romero D, Kopka CJ, Karim SA, Abu-Raddad LJ, Almeida G, et al. A multinational Delphi consensus to end the COVID-19 public health threat. Nature. 2022;611(7935):332–45.

11. Lazarus JV, Safreed-Harmon K, Kamarulzaman A, Anderson J, Leite RB, Behrens G, et al. Consensus statement on the role of health systems in advancing the long-term well-being of people living with HIV. Nature Communications. 2021;12(1):4450.

12. Winter RJ, Sheehan Y, Papaluca T, Macdonald GA, Rowland J, Colman A, et al. Consensus recommendations on the management of hepatitis C in Australia’s prisons. Med J Aust. 2023;218(5):231–7.

13. Hedrich D, Alves P, Farrell M, Stöver H, Møller L, Mayet S. The effectiveness of opioid maintenance treatment in prison settings: a systematic review. Addiction. 2012;107(3):501–17.

14. Larney S. Does opioid substitution treatment in prisons reduce injecting-related HIV risk behaviours? A systematic review. Addiction. 2010;105(2):216–23.

15. Cates L, Brown AR. Medications for opioid use disorder during incarceration and post-release outcomes. Health & Justice. 2023;11(1).

16. Moore KE, Roberts W, Reid HH, Smith KMZ, Oberleitner LMS, McKee SA. Effectiveness of medication assisted treatment for opioid use in prison and jail settings: A meta-analysis and systematic review. J Subst Abuse Treat. 2019;99:32–43.

17. Peters RH, Greenbaum PE, Steinberg ML, Carter CR, Ortiz MM, Fry BC, et al. Effectiveness of screening instruments in detecting substance use disorders among prisoners. Journal of Substance Abuse Treatment. 2000;18(4):349–58.

18. Wolff N, Shi J. Screening for Substance Use Disorder Among Incarcerated Men with the Alcohol, Smoking, Substance Involvement Screening Test (ASSIST): A Comparative Analysis of Computer-Administered and Interviewer-Administered Modalities. J Subst Abuse Treat. 2015;53:22–32.

19. Ray B, Victor G, Cason R, Hamameh N, Kubiak S, Zettner C, et al. Developing a cascade of care for opioid use disorder among individuals in jail. J Subst Abuse Treat. 2022;138:108751.

20. Darke S, Larney S, Farrell M. Yes, people can die from opiate withdrawal. Addiction. 2017;112(2):199–200.

21. Stewart AC, Cossar RD, Wilkinson AL, Quinn B, Dietze P, Walker S, et al. The Prison and Transition Health (PATH) cohort study: Prevalence of health, social, and crime characteristics after release from prison for men reporting a history of injecting drug use in Victoria, Australia. Drug and Alcohol Dependence. 2021;227:108970.

22. Curtis M, Winter RJ, Dietze P, Wilkinson AL, Cossar RD, Stewart AC, et al. High rates of resumption of injecting drug use following release from prison among men who injected drugs before imprisonment. Addiction. 2022;117(11):2887–98.

23. Binswanger IA, Stern MF, Deyo RA, Heagerty PJ, Cheadle A, Elmore JG, et al. Release from Prison — A High Risk of Death for Former Inmates. New England Journal of Medicine. 2007;356(2):157–65.

24. Borschmann R, Borschmann R, Keen C, Spittal MJ, Preen D, Pirkis J, et al. Rates and causes of death after release from incarceration among 1L471L526 people in eight high-income and middle-income countries: an individual participant data meta-analysis. The Lancet. 2024;403(10438):1779–88.

25. Curtis M, Larney S, Higgs P, Cossar RD, Winter RJ, Stewart AC, et al. Initiation of Medications for Opioid Use Disorder Shortly Before Release From Prison to Promote Treatment Retention: Strong Evidence but Compromised Policy. Journal of Addiction Medicine. 2021;15(6).

26. Kinlock TW, Gordon MS, Schwartz RP, Fitzgerald TT, O’Grady KE. A randomized clinical trial of methadone maintenance for prisoners: Results at 12 months postrelease. Journal of Substance Abuse Treatment. 2009;37(3):277–85.

27. Wright NM, Sheard L, Adams CE, Rushforth BJ, Harrison W, Bound N, et al. Comparison of methadone and buprenorphine for opiate detoxification (LEEDS trial): a randomised controlled trial. British Journal of General Practice. 2011;61(593):e772–e80.

28. Dunlop AJ, White B, Roberts J, Cretikos M, Attalla D, Ling R, et al. Treatment of opioid dependence with depot buprenorphine (CAM2038) in custodial settings. Addiction. 2022;117(2):382–91.

29. Tracy MC, Thompson R, Muscat DM, Bonner C, Hoffmann T, McCaffery K, et al. Implementing shared decision-making in Australia. Zeitschrift für Evidenz, Fortbildung und Qualität im Gesundheitswesen. 2022;171:15–21.

30. Windle E, Tee H, Sabitova A, Jovanovic N, Priebe S, Carr C. Association of Patient Treatment Preference With Dropout and Clinical Outcomes in Adult Psychosocial Mental Health Interventions: A Systematic Review and Meta-analysis. JAMA Psychiatry. 2020;77(3):294–302.

31. Puglisi LB, Bedell PS, Steiner A, Wang EA. Medications for Opioid Use Disorder Among Incarcerated Individuals: a Review of the Literature and Focus on Patient Preference. Current Addiction Reports. 2019;6(4):365–73.

32. Kaplowitz E, Truong AQ, Berk J, Martin RA, Clarke JG, Wieck M, et al. Treatment preference for opioid use disorder among people who are incarcerated. J Subst Abuse Treat. 2022;137:108690.

33. Larance B, Degenhardt L, Lintzeris N, Winstock A, Mattick R. Definitions related to the use of pharmaceutical opioids: Extramedical use, diversion, non-adherence and aberrant medication-related behaviours. Drug and Alcohol Review. 2011;30(3):236–45.

34. Bi-Mohammed Z, Wright NM, Hearty P, King N, Gavin H. Prescription opioid abuse in prison settings: A systematic review of prevalence, practice and treatment responses. Drug Alcohol Depend. 2017;171:122–31.

35. White N, Ali R, Larance B, Zador D, Mattick RP, Degenhardt L. The extramedical use and diversion of opioid substitution medications and other medications in prison settings in Australia following the introduction of buprenorphine–naloxone film. Drug and Alcohol Review. 2016;35(1):76–82.

36. Ling R, White B, Roberts J, Cretikos M, Howard MV, Haber PS, et al. Depot buprenorphine as an opioid agonist therapy in New South Wales correctional centres: a costing model. BMC Health Services Research. 2022;22(1).

37. Wright N, Hard J, Fearns C, Gilman M, Littlewood R, Clegg R, et al. OUD Care Service Improvement with Prolonged-release Buprenorphine in Prisons: Cost Estimation Analysis. ClinicoEconomics and Outcomes Research. 2020;12(null):499–504.

38. Roberts J, White B, Attalla D, Ward S, Dunlop AJ. Rapid upscale of depot buprenorphine (CAM2038) in custodial settings during the early COVID-19 pandemic in New South Wales, Australia. Addiction. 2021;116(2):426–7.

39. Marmel A, Bozinoff N. Punitive discontinuation of opioid agonist therapy during incarceration. Int J Prison Health. 2020;16(4):337–42.

40. Brinkley-Rubinstein L, McKenzie M, Macmadu A, Larney S, Zaller N, Dauria E, et al. A randomized, open label trial of methadone continuation versus forced withdrawal in a combined US prison and jail: Findings at 12 months post-release. Drug Alcohol Depend. 2018;184:57–63.

41. Rich JD, McKenzie M, Larney S, Wong JB, Tran L, Clarke J, et al. Methadone continuation versus forced withdrawal on incarceration in a combined US prison and jail: a randomised, open-label trial. Lancet. 2015;386(9991):350–9.

42. Maradiaga JA, Nahvi S, Cunningham CO, Sanchez J, Fox AD. “I Kicked the Hard Way. I Got Incarcerated.” Withdrawal from Methadone During Incarceration and Subsequent Aversion to Medication Assisted Treatments. J Subst Abuse Treat. 2016;62:49–54.

43. Cooper JA, Onyeka I, Cardwell C, Paterson E, Kirk R, O’Reilly D, et al. Record linkage studies of drug-related deaths among adults who were released from prison to the community: a scoping review. BMC Public Health. 2023;23(1).

44. Forsyth SJ, Carroll M, Lennox N, Kinner SA. Incidence and risk factors for mortality after release from prison in Australia: a prospective cohort study. Addiction. 2018;113(5):937–45.

45. Curtis M, Wilkinson AL, Dietze P, Stewart AC, Kinner SA, Cossar RD, et al. Prospective study of retention in opioid agonist treatment and contact with emergency healthcare following release from prisons in Victoria, Australia. Emerg Med J. 2023;40(5):347–54.

46. Curtis M, Wilkinson AL, Dietze P, Stewart AC, Kinner SA, Winter RJ, et al. Is use of opioid agonist treatment associated with broader primary healthcare use among men with recent injecting drug use histories following release from prison? A prospective cohort study. Harm Reduct J. 2023;20(1):42.

47. Curtis M, Dietze P, Wilkinson AL, Agius PA, Stewart AC, Cossar RD, et al. Discontinuation of opioid agonist treatment following release from prison in a cohort of men who injected drugs prior to imprisonment in Victoria, Australia: A discrete-time survival analysis. Drug and Alcohol Dependence. 2023;242:109730.

48. Larney S, Toson B, Burns L, Dolan K. Effect of prison-based opioid substitution treatment and post-release retention in treatment on risk of re-incarceration. Addiction. 2012;107(2):372–80.

49. Krulic T, Brown G, Bourne A. A Scoping Review of Peer Navigation Programs for People Living with HIV: Form, Function and Effects. AIDS and Behavior. 2022;26(12):4034–54.

50. McBrien KA, Ivers N, Barnieh L, Bailey JJ, Lorenzetti DL, Nicholas D, et al. Patient navigators for people with chronic disease: A systematic review. PLOS ONE. 2018;13(2):e0191980.

51. Sullivan E, Zeki R, Ward S, Sherwood J, Remond M, Chang S, et al. Effects of the Connections program on return-to-custody, mortality and treatment uptake among people with a history of opioid use: Retrospective cohort study in an Australian prison system. Addiction. 2024;119(1):169–79.

52. Schwartz RP, Kelly SM, Mitchell SG, O’Grady KE, Sharma A, Jaffe JH. Methadone treatment of arrestees: A randomized clinical trial. Drug Alcohol Depend. 2020;206:107680.

53. Mitchell SG, Harmon-Darrow C, Lertch E, Monico LB, Kelly SM, Sorensen JL, et al. Views of barriers and facilitators to continuing methadone treatment upon release from jail among people receiving patient navigation services. Journal of Substance Abuse Treatment. 2021;127:108351.

54. Enich M, Treitler P, Swarbrick M, Belsky L, Hillis M, Crystal S. Peer Health Navigation Experiences Before and After Prison Release Among People With Opioid Use Disorder. Psychiatric Services. 2023;74(7):737–45.

55. Tillson M, Fallin-Bennett A, Staton M. Providing peer navigation services to women with a history of opioid misuse pre- and post-release from jail: A program description. Journal of Clinical and Translational Science. 2022;6(1):1–23.

56. Kendall S, Redshaw S, Ward S, Wayland S, Sullivan E. Systematic review of qualitative evaluations of reentry programs addressing problematic drug use and mental health disorders amongst people transitioning from prison to communities. Health Justice. 2018;6(1):4.

57. Bird SM, McAuley A, Munro A, Hutchinson SJ, Taylor A. Prison-based prescriptions aid Scotland’s National Naloxone Programme. The Lancet. 2017;389(10073):1005–6.

58. Bird SM, McAuley A, Perry S, Hunter C. Effectiveness of Scotland’s National Naloxone Programme for reducing opioid-related deaths: a before (2006-10) versus after (2011-13) comparison. Addiction. 2016;111(5):883–91.

59. Curtis M, Dietze P, Aitken C, Kirwan A, Kinner SA, Butler T, et al. Acceptability of prison-based take-home naloxone programmes among a cohort of incarcerated men with a history of regular injecting drug use. Harm Reduction Journal. 2018;15(1):48.

60. Moradmand-Badie B, Tran L, Oikarainen N, Degenhardt L, Nielsen S, Roberts J, et al. Feasibility and acceptability of take-home naloxone for people released from prison in New South Wales, Australia. Drug Alcohol Rev. 2021;40(1):98–108.

61. Australian Bureau of Statistics. Prisoners in Australia 2023 [Available from: https://www.abs.gov.au/statistics/people/crime-and-justice/prisoners-australia/latest-release#data-downloads.

62. Campbell MA, Hunt J, Scrimgeour DJ, Davey M, Jones V. Contribution of Aboriginal Community-Controlled Health Services to improving Aboriginal health: an evidence review. Australian Health Review. 2018;42(2):218–26.

63. 63. Declaration on the Rights of Indigenous People, (2007).

64. Sivak L, Cantley L, Kelly J, Reilly R, Hawke K, Mott K, et al. Model of Care for Aboriginal Prisoner Health and Wellbeing for South Australia – Final Report. Adelaide, South Australia: Wardliparingga Aboriginal Health Research Unit: SAHMRI; 2017.

65. Tongs J, Chatfield H, Arabena K. The Winnunga Nimmityjah Aboriginal Health Service Holistic Health Care for Prison Model. Aboriginal and Islander Health Worker Journal. 2007;31(6):6–8.

66. Pettit S, Simpson P, Jones J, Williams M, Islam MM, Parkinson A, et al. Holistic primary health care for Aboriginal and Torres Strait Islander prisoners: exploring the role of Aboriginal Community Controlled Health Organisations. Australian and New Zealand Journal of Public Health. 2019;43(6):538–43.

67. Freeburn B, Loggins S, Lee KSK, Conigrave KM. Coming of age: 21 years of providing opioid substitution treatment within an Aboriginal community-controlled primary health service. Drug Alcohol Rev. 2022;41(1):260–4.

68. Australian Institute of Health and Welfare (AIHW). The health of people in Australia’s prisons 2022. Canberra: Australian Government; 2022 8 January 2023.

69. Steely Smith MK, Wilson SH, Zielinski MJ. An integrative literature review of substance use treatment service need and provision to pregnant and postpartum populations in carceral settings. Womens Health (Lond). 2023;19:17455057221147802.

70. Winklbaur B, Kopf N, Ebner N, Jung E, Thau K, Fischer G. Treating pregnant women dependent on opioids is not the same as treating pregnancy and opioid dependence: a knowledge synthesis for better treatment for women and neonates. Addiction. 2008;103(9):1429–40.

71. Krans EE, Kim JY, Chen Q, Rothenberger SD, James AE, Kelley D, et al. Outcomes associated with the use of medications for opioid use disorder during pregnancy. Addiction. 2021;116(12):3504–14.

72. Terplan M, Laird HJ, Hand DJ, Wright TE, Premkumar A, Martin CE, et al. Opioid Detoxification During Pregnancy: A Systematic Review. Obstet Gynecol. 2018;131(5):803–14.

73. Ordean A, Tubman-Broeren M. Safety and Efficacy of Buprenorphine-Naloxone in Pregnancy: A Systematic Review of the Literature. Pathophysiology. 2023;30(1):27–36.

74. Peeler M, Fiscella K, Terplan M, Sufrin C. Best Practices for Pregnant Incarcerated Women With Opioid Use Disorder. J Correct Health Care. 2019;25(1):4–14.

75. Kim SB, White B, Roberts J, Day CA. Substance use among pregnant women in NSW prisons. International Journal of Drug Policy. 2023;122:104256.

76. Australian Institute of Health and Welfare (AIHW). National Opioid Pharmacotherapy Statistics Annual Data Collection 2022. 2023.

77. Grella CE, Ostile E, Scott CK, Dennis M, Carnavale J. A Scoping Review of Barriers and Facilitators to Implementation of Medications for Treatment of Opioid Use Disorder within the Criminal Justice System. International Journal of Drug Policy. 2020;81:102768.

