## Appendix 1 for "National consensus statement on opioid agonist treatment in custodial settings"

### Appendices

Appendix 1: Consensus Statement Expert Panel

| **Name** | **Affiliation** | **State/**  **Territory** |
| --- | --- | --- |
| Adrian Dunlop | Director and Senior Staff Specialist, Drug and Alcohol Clinical Services, Hunter New England Local Health District, New South Wales; President of the Australasian Chapter of Addiction Medicine, Royal Australasian College of Physicians | NSW |
| Andrew Wiley | Director, SA Prison Health Service, Nurse | SA |
| Bianca Davidde | Addiction Medicine Physician, Drug and Alcohol Services, DASSA | SA |
| Christine Watson | Director of the Addiction Medical Services, Northern Territory | NT |
| David Onu | Forensic Medical Specialist & General Practitioner, Statewide Specialty Director, Correctional Health Services | TAS |
| Ele Morrison | Director of Advocacy, Australian Injecting & Illicit Drug Users League | VIC |
| Jeremy Hayllar | Clinical Director of the Alcohol and Drug Service of Metro North Mental Health - Alcohol and Drug Service | QLD |
| Jocelyn Chan | Addiction Medicine Registrar & Public Health Physician, Western Health Drug Health Services | VIC |
| Katerina Lagios | Sexual Health Physician and Clinical Director Population Health, Justice Health & Forensic Mental Health Network, New South Wales | NSW |
| Kevin Fontana | Medical Services Director, Corrective Services, Department of Justice, Western Australia | WA |
| Mark Stoove | Head of Public Health, Burnet Institute | VIC |
| Peter Thompson | Co-Clinical Director Drug & Alcohol, Justice Health and Forensic Mental Health Network, New South Wales | NSW |
| Rebecca Li | Director of Clinical Services within Canberra Health Services | ACT |
| Rebecca Winter | Deputy Head Justice Health Group, Burnet Institute | VIC |
| Shalini Arunogiri | Clinical Director, Statewide Centre for Addiction and Mental Health, Turning Point | VIC |
| Suzanne Nielsen | Deputy Director of the Monash Addiction Research Centre, Monash University; Pharmacist; President-Elect Australasian Professional Society on Alcohol and Other Drugs | VIC |
| Thileepan Naren | Addiction Medicine Physician, Western Health | VIC |
| Tom Turnbull | Medical Director, Prison Health Service, South Australia; Chair of the Royal Australasian College of General Practitioners Specific Interest Group for Custodial Health | SA |
